# Positivity of SARS-CoV-2, by RT-PCR among workers of a Public Hospital in the city of Santos, SP, Brazil 2020

**DOI:** 10.1101/2020.06.30.20143529

**Authors:** Marcos Montani Caseiro, Monica Mazzurana, Bruno Barreiro, Estela Capelas Barbosa, Antônio Pires Barbosa

## Abstract

**Background:** Infection with SARS-CoV-2, associated with severe respiratory symptoms, currently called COVID-19, represents one of the greatest health challenges of this century. Health professionals, as well as hospital workers, are the most vulnerable population for acquiring SARS-CoV-2 due to their greater proximity to sick patients, especially in specialized hospitals for the treatment of these patients.

**Study Design:** We ran a questionnaire by all the employees of a Public Service Hospital specialised in COVID-19 treatment (1307 employees), in the municipality of Santos, São Paulo, Brazil, who presented some symptoms compatible with COVID-19 and positive results by RT-PCR, in Oropharynx Swab collection. The questionnaire collected socio-demographic information, as well as data related to the work areas where they were divided into two groups: Employees at higher risk or those who directly deal with patients (914 employees or 69.9%) and lower risk for employees in the administrative area (393 employees or 30.1%).

**Results:** 211 (16.1%) of the Hospital’s employees had to stop their work-related activities due to presenting positive RT-PCR. There were 39 (9.9%) positives in low risk areas and 172 (18.8%) positives in employees in high risk areas. Within the latter group, Nurses, Nursing Assistants and Doctors were the most frequently infected professionals. Regarding the symptoms of professionals positively diagnosed based on RT-PCR, the most frequent symptoms were body pain (83.4%); headache (80.6%); fever (57.8%) and dry cough (53.1%).

**Conclusion:** A high proportion of workers in COVID-19 specialized hospitals was infected by the virus, despite all the protective measures adopted, showing the high transmission capacity of this virus. Stricter individual protection measures among employees must be adopted.

## 1. Introduction

The first reports from the virus started with the identification of patients with an atypical respiratory syndrome in Wuhan, in the Chinese province of Hubei, in early December (Yan et al. 2020; Shi et al. 2020). On December 27th, 2019, the Chinese Disease Control Center (CDC) notified atypical pneumonia cases with unusual evolution and a few days later, on December 31st, Wuhan’s epidemiological surveillance issued an alert to the Chinese CDC, which promptly started investigating the cases, their etiology and their probable origin. A day later, on January 1st, 2020, CDC investigators related the cases to the Huanan fish market, which was promptly closed (Zhou et al. 2020); the causative agent of the disease was identified and initially called the new Coronavirus - (nCoV-2019), being later renamed Severe Acute Respiratory Syndrome Coronavirus-2, (SARS-CoV-2) and the disease later named - Coronavirus 2019 Disease (COVID-19) (Zhu et al. 2020), see figure 1.

**Figure 1.**
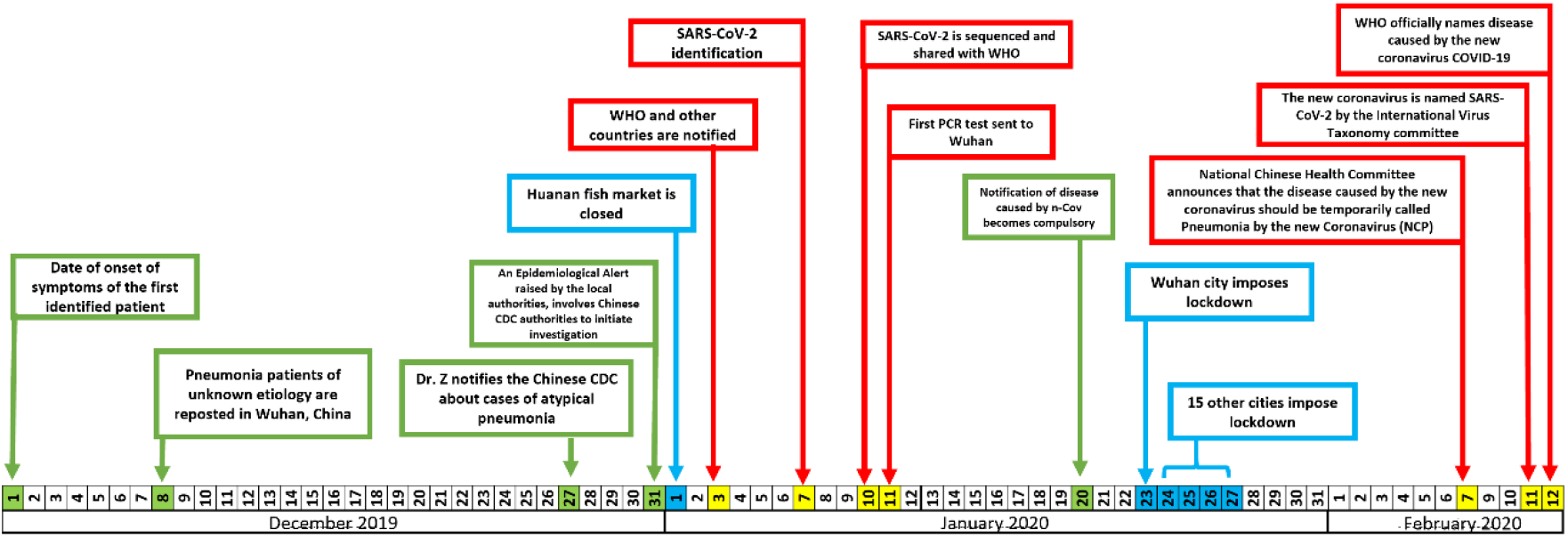
Timeline with the most relevant facts associated with the emergence of Severe Acute Respiratory Syndrome by Coronavirus 2, (SARS-CoV2 / COVD-19)

The current number of people infected confirmed in the world as of 06 June 2020 is 7,936,874 and 433,959 deaths. In Brazil, there are 867,624 cases and 43,332 reported deaths. The municipality of Santos is located in the coastal region of the State of São Paulo. It is the locus of the largest port in Latin America and it has a current population of 443,000 inhabitants. In the municipality of Santos, the first reported case was described retrospectively dating back to 05 Feb 2020; and currently the number of confirmed patients is 6354 and 222 deaths.

## 2. Aims

To determine the prevalence of positive RT-PCR-SARS-CoV-2 among healthcare workers at Hospital Guilherme Álvaro, Santos, SP / Brazil and their sociodemographic and clinical characteristics.

## 3. Study Design

We analyzed all employees of Hospital Guilherme Álvaro who were removed from their work activities from February to June 2020, as a result of being diagnosed with COVID-19, using an Oropharynx Swab. They were stratified according to sociodemographic characteristics, reported symptoms, work areas, second or minor contact with patients (Assistance Area and Administrative Area / Service) and type of healthcare professional. RT-PCR tests were performed using the Charité protocol (Corman et al. 2020); based on reverse transcription followed by real-time Polymerase chain reaction of the three Coronavirus -SARS-CoV-2 genes (Gene E, RdRP and N). All data were collected through a semi-structured questionnaire and sent out via Google Forms® platform. The data was initially condensed in an Excel-Microsoft® spreadsheet, and later analyzed by the SPSS-IBM® statistical package - version 25, 2017.

The data was presented in tables, including absolute numbers and relative proportions for the different variables, using Fisher’s ex(Corman et al. 2020)act test. This study was submitted to the Ethics and Research Committee of Hospital Guilherme Álvaro, and approved under protocol number 4.045.803.

## 4. Results

From a total of 1307 healthcare workers in the different areas of the hospital, 211 (16.1%) of hospital employees had to remain on leave for at least 14 days due to the diagnosis of COVID-19, graph 1.

**Graph 1.**
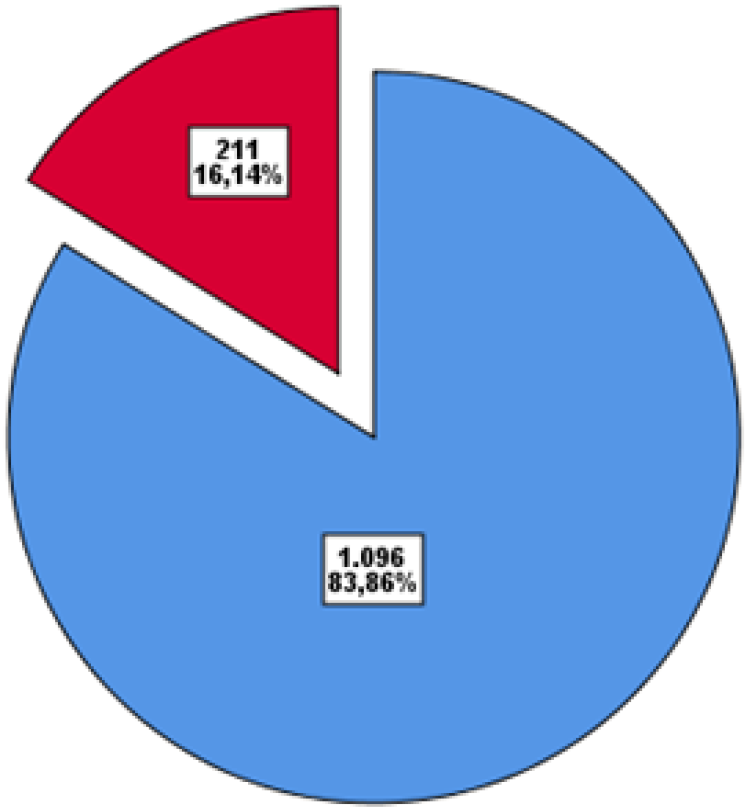
Absolute and relative frequencies of employees with RT-PCR-SARS-CoV-2, away from work at Hospital Guilherme Álvaro, Santos, SP, Brazil, between Feb and Jun 2020.

The different sociodemographic characteristics of these, as well as means of transportation to get to work and commuting times were stratified by work area considered to be of greater or lesser risk and can be seen in Table 1.

**Table 1.** Absolute and relative frequencies of sociodemographic variables of employees at Hospital Guilherme Álvaro, 2020.

|  |  | Administrative / Services | Clinical Care | Total |
| --- | --- | --- | --- | --- |
| GENDER | Female | 266 (27,9%) | 687 (72,1%) | 953 (72,7%) |
|  | Male | 127 (35,9%) | 227 (64,1%) | 354 (27,3%) |
| AGE | Mean | 45.8 | 44.8 | 45.1 |
|  | Median | 47 | 46 | 46 |
|  | Mode | 54 | 47 | 54 |
| ETHNICITY | White | 193 (49,1%) | 587 (64,2%) | 780 (59,7%) |
|  | Mixed | 162 (41,2%) | 238 (26%) | 400 (30,6%) |
|  | Black | 31 (7,9%) | 75 (8,2%) | 106 (8,1%) |
|  | Asian | 5 (1,3%) | 13 (1,4%) | 18 (1,4%) |
|  | Indigenous | 2 (0,5%) | 1 (0,1%) | 3 (0,2%) |
| MEANS OF TRANSPORT TO WORK | Car | 106 (27%) | 524 (57,3%) | 630 (48,2%) |
|  | Bus/Van | 190 (48,3%) | 207 (22,6%) | 397 (30,4%) |
|  | Walking | 37 (9,4%) | 81 (8,9%) | 118 (9,0%) |
|  | Motorcycle | 33 (8,4%) | 76 (8,3%) | 109 (8,3%) |
|  | Bicycle | 27 (6,9%) | 26 (2,8%) | 53 (4,1%) |
| COMMUTING TIME | <30 minutes | 147 (37,4%) | 526 (57,5%) | 673 (51,5%) |
|  | 30 - 60 ' | 156 (39,7%) | 256 (28 %) | 412 (31,5%) |
|  | 60 e 120 ' | 72 (18,3%) | 111 (12,1%) | 183 (14%) |
|  | > 120' | 18 (4,6%) | 21 (2,3%) | 39 (3,0%) |
| LEVEL OF SCHOOLING | Incomplete Primary | 34 (8,7%) | 12 (1,3%) | 46 (3,5%) |
|  | Complete Primary | 25 (6,4%) | 16 (1,8%) | 41 (3,1%) |
|  | Incomplete Secondary | 21 (5,3%) | 11 (1,2%) | 32 (2,4%) |
|  | Complete Secondary | 136 (34,6%) | 205 (22,4%) | 341 (26,1%) |
|  | Incomplete Higher Education | 43 (10,9%) | 77 (8,4%) | 120 (9,2%) |
|  | Complete Higher Education | 83 (21,1%) | 256 (28%) | 339 (25,9%) |
|  | Post-graduate level | 43 (10,9%) | 255 (27,9%) | 298 (22,8%) |
|  | Masters | 7 (1,8%) | 57 (6,2%) | 64 (4,9%) |
|  | PhD | 1 (0,3%) | 25 (2,7%) | 26 (2,0%) |

The patients were initially stratified by work area, according to the greater or lesser risk, clinical care and administrative / services respectively. The prevalence of a positive Swab RT-PCR-SARS-CoV-2 was 2.1 times higher in clinical care area than in the administrative area, 172 (18.8%) and 39 (9.9%) respectively (Table 2).

**Table 2.**
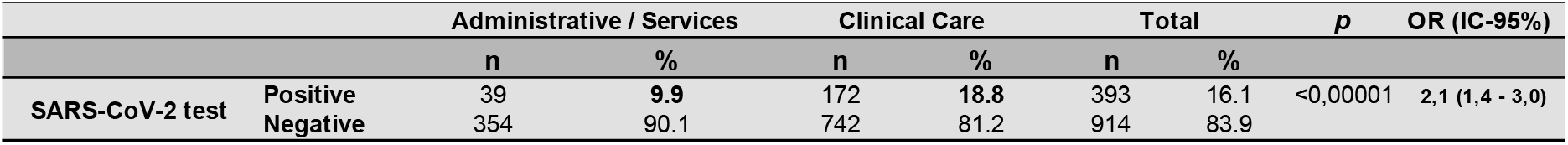
Absolute and relative frequencies of the prevalence of Swab RT-PCR-SARS-CoV-2, stratified by the different work areas of the employees of Hospital Guilherme Álvaro, 2020.

Subsequently, employees with a positive diagnosis were stratified according to gender, age (greater than or less than 60 years old), and the professional’s specialty in relation to their activity with patients. Positivity was more frequent among women; employees under the age of 60 years, and the nursing group (technicians and assistants) (Table 3).

**Table 3.**
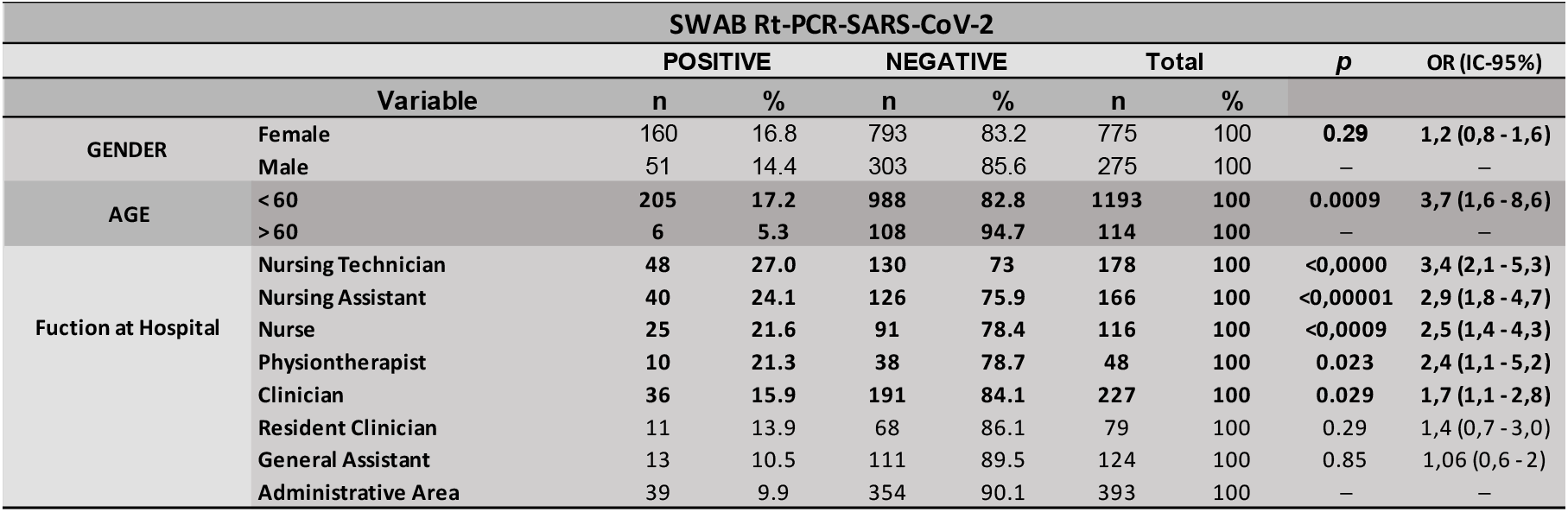
Absolute and relative frequencies, *p* and Odds Ratio (OR) -IC-95%, of employees stratified by the Swab RT-PCR-SARS-CoV-2 result, according to gender, age and professional activity, Hospital Guilherme Álvaro, 2020.

Finally, the main clinical complaints associated with patients diagnosed with COVID-19 were evaluated. In short, we found that body pain was the most frequent symptom, being reported by 83.4% of patients, and the fever and cough, which are most often associated with the diagnosis of COVID-19, were present in 57.8% and 53.1%, respectively, only (Table 4).

**Table 4.**
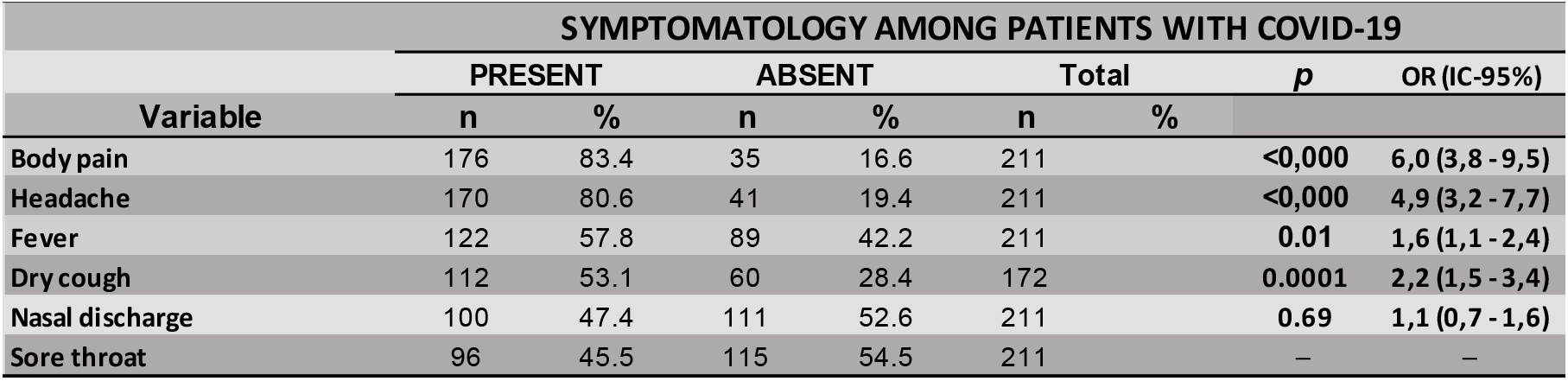
Absolute and relative frequencies, *p* and Odds Ratio (OR) -IC-95%, of the main symptoms among patients with COVID-19 at Hospital Guilherme Álvaro, 2020.

## Discussion

This study represents the first step in tackling the COVID-19 pandemic, which began in determining the prevalence of Hospital employees in different areas of work. 211 of 1307 (16.1%) employees were removed from their usual activities, due to the presence of symptoms and positive diagnosis based on oropharynx swab for RT-PCR detection. The prevalence of infection among workers in the high-risk clinical care area was 18.8% (172/393) and in the administrative and services area, where there is less contact with patients, therefore were considered low risk, we found 9.9% (39/393); with an OR of 2.1 (1.4 - 3.0 - 95% CI) for individuals in the high risk area compared to the low risk. Other studies using similar methodology, carried out through the collection of RT-PCR, showed an infection rate close to ours. In the United Kingdom, among 1533 symptomatic healthcare workers studied in a tertiary hospital 282 (18%) tested positive (Keeley et al. 2020); these authors also concluded that early identification in symptomatic patients is of fundamental importance to avoid the transmission of the disease.

Results of studies using antibody tests through serology have shown inferior results among these same professionals; Korth in Germany in a study among healthcare professionals found a 1.6% prevalence among 316 workers, and after stratifying the cases by areas of high risk, thes prevalence rose to 5.4% (Korth et al. 2020); in Santa Clara, USA, a study with approximately 3,500 healthcare workers, the prevalence was 1.5% (Zwald et al. 2020). There was no statistically significant difference in positive diagnosis in relation to gender, however, there was a statistically significant difference between workers aged 60 years or less (table 3); this certainly reflects the hospital’s policy of removing the most vulnerable professionals from the areas at greatest risk (Giannis et al. 2020).When comparing the characteristics of individuals who had a positive Swab RT-PCR test, in relation to the health professional’s role according to their workplace, a higher OR was observed in workers from higher risk areas (table 3); thus the odds ratio was larger than one for healthcare professionals with greater contact with patients, such as a nursing technician - OR-3,4; nursing assistant - OR = 2.9; nurses - OR-2.5; physiotherapists - OR-2,4 and clinicians - OR-1,7; all statistically significant. Among health professionals with positive diagnoses who were promptly removed from their work activities at the hospital, we investigated the main symptoms reported, and what draws attention is the fact that, even though most studies show the occurrence of fever and cough as predominant symptoms (Guan et al. 2020; Huang et al. 2020; Lescure et al. 2020), in symptomatic patients in the present study, the predominant symptom was muscle pain followed by headache (table 4); fever was only the third symptom reported in 57.8% of patients, showing to some extent the uselessness of temperature measurement in many locations as a measure of early detection of patients with COVID-19. In our view, symptoms such as muscle pain and headache, even if isolated, should raise the suspicion of infection, at least in this pandemic period. In addition, asymptomatic infections were observed in other studies in up to 50% of cases (Day 2020; Mizumoto et al. 2020; Shim et al. 2020), with the transmissibility period occurring even before the onset of symptoms (He et al. 2020). Therefore, we have reviewed the measures of patient evaluation as possible symptoms that could lead to sick leave.

In conclusion, the prevalence of COVID-19 among healthcare professionals in a tertiary hospital in the municipality of Santos, SP, Brazil was 16.1%. Despite the implementation of training for all health professionals regarding measures and adequate handling of personal protective equipment, continued training measures for healthcare professionals in areas of treatment of patients with COVID-19 must be adopted, as well as removal of the most vulnerable professionals from the areas of greatest risk. The next stage of this study will be carrying out serology in all professionals employed by the hospital, with a follow-up study of seroconversion among the negative and a drop in antibody titers among the positive, every three months for 2 years.

## Data Availability

http://saude.sp.gov.br/coordenadoria-de-recursos-humanos/areas-da-crh/qualidade-de-vida-do-trabalhador-da-saude/programa-de-preparacao-para-a-aposentadoria/o-programa-nas-unidades/hospital-guilherme-alvaro-em-santos

